## Supplementary material for "Unified platform for genetic and serological detection of COVID-19 with single-molecule technology": Table S4

**Table S4. Clinical features of serum samples tested from patients with active disease.**

|  | **Symptoms** | **Days since symptom onset** | **disease severity** | **IgM (median)** | **IgG (median)** |
| --- | --- | --- | --- | --- | --- |
| P26 | symptomatic | 4 | moderate | 830 ± 45 | 456 ± 76 |
| P24 | symptomatic | 9 | moderate | 673 ± 85 | 225 ± 73 |
| P22 | symptomatic | 9 | mild | 497 ± 127 | 464 ± 115 |
| P23 | symptomatic | 6 | mild | 353 ± 78 | 238 ± 101 |
| P25 | asymptomatic | 2 days after positive qPCR | mild | 164 ± 17 | 75 ± 10 |
