## Supplementary material for "Unified platform for genetic and serological detection of COVID-19 with single-molecule technology": Table S3

| **Sample** | 7 | 23 | 5 | 3 | 17 | 22 | 2 | 12 | 19 | 13 | 6 |
| --- | --- | --- | --- | --- | --- | --- | --- | --- | --- | --- | --- |
| **Ct (GeneE)** | 32.8 | 17.7 | 31.9 | 32.5 | 23.4 | 33.6 | 27.2 | 20.3 | 23.4 | 24.3 | 11.9 |
| **Single-molecule (median)** | 69* | 73* | 78* | 87.5* | 101.5* | 163 | 165 | 190.5 | 199.5 | 206 | 1070.5 |

**Table S3.** **Ct values (gene E) and single-molecule scores (median of number of spots/ FOV) for positive swab samples presented in figure 1D.**

* Single-molecule signal is below classification cutoff.
