## Supplementary figures and images for "Unified platform for genetic and serological detection of COVID-19 with single-molecule technology"

### S1 figure

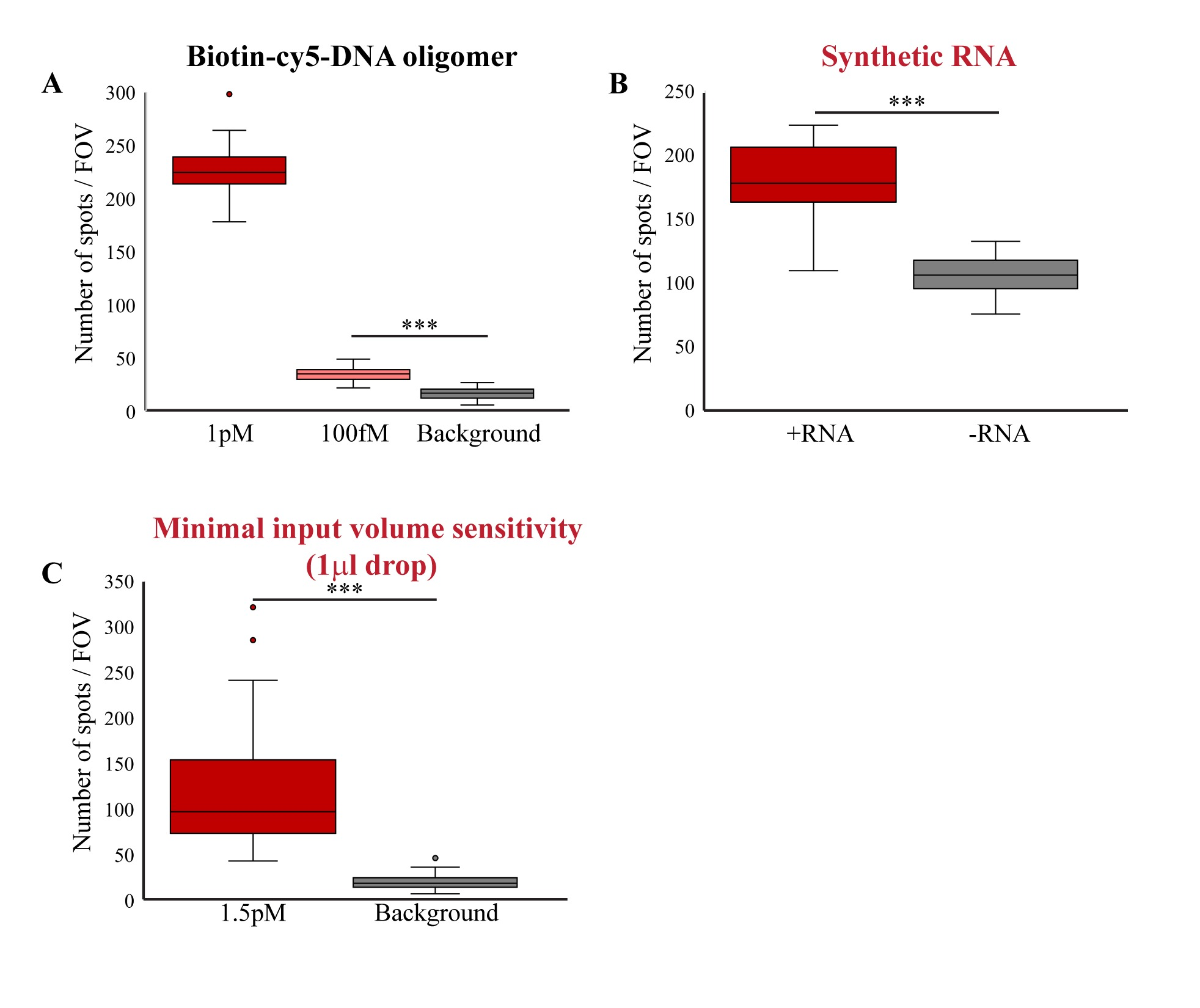

### S2 figure

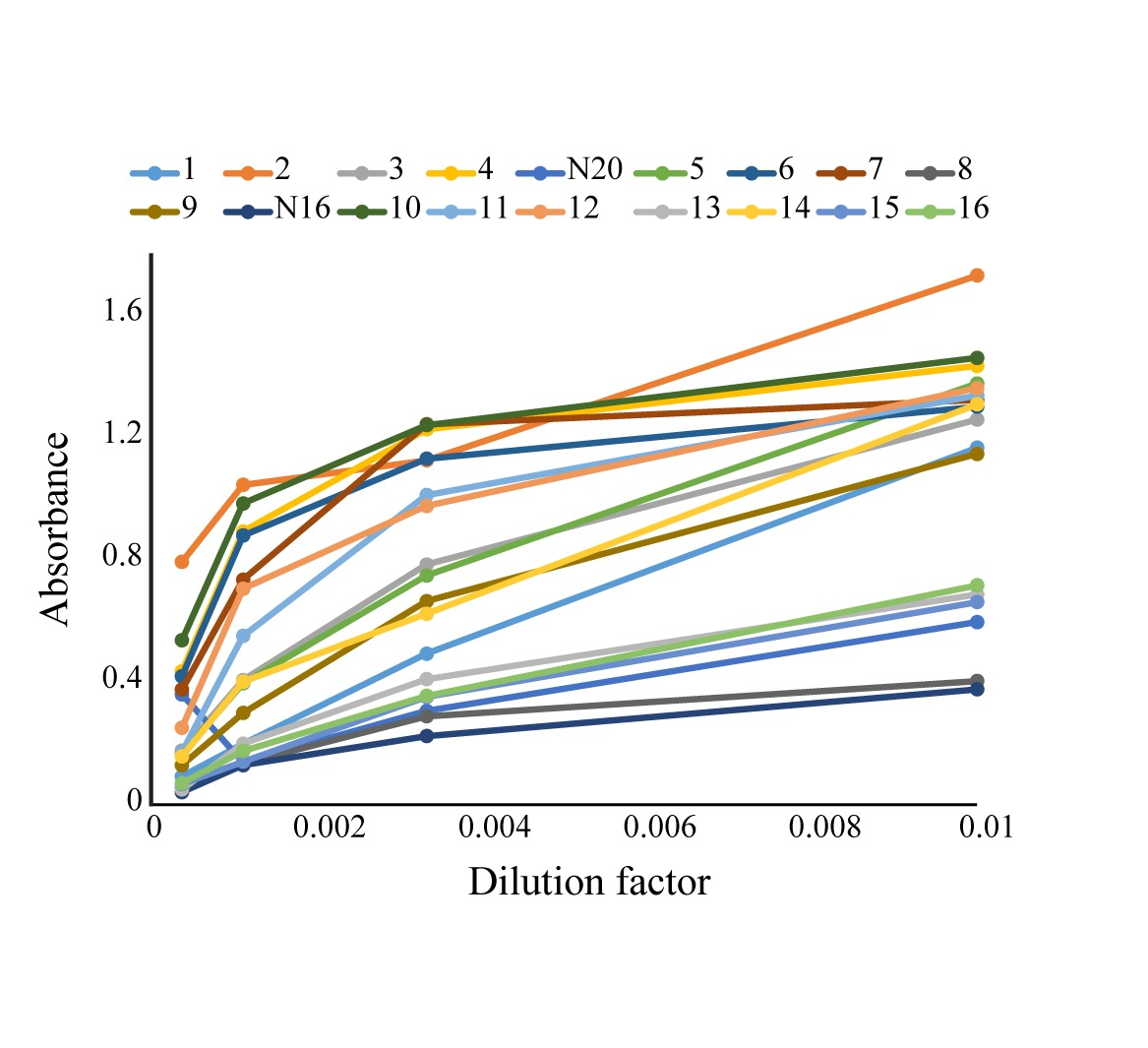
